# Clinical efficacy of Early Administration of Convalescent Plasma among COVID-19 Cases in Egypt

**DOI:** 10.1101/2021.03.30.21254031

**Authors:** Noha Asem, Hossam Hosny Massoud, Ihab Serag, Mohamed Hassany, Gehan ElAssal, Akram Abdelbary, Marwa Mohsen, Amin Abdel Baki, Samy Zaky, Wagdy Amin, Ehab Kamal, Hamdy Ibrahem, Ahmed Said Abdel Mohsen, Mohamed Ahmed Ali, Nancy Elgendy, Mohamed Elbadry, Salwa Hassan Ahmed, Ahmed Naguib Nassif shenouda, Mohamed Abdelhamed Fathy, Hala Zaid

**Author notes:** Corresponding Author Noha Asem, Cairo University, Ministry of Health and Population, Postal code: 11311, Cairo, Egypt. Last academic degree. Doctorate degree (Noha Asem, Hossam Hosny Massoud, Mohamed Hassany, Gehan ElAssal, Akram Abdelbary, Amin Abdel Baki, Samy Zaky, Wagdy Amin, Ehab Kamal, Hamdy Ibrahem, Ahmed Said Abdel Mohsen, Mohamed Ahmed Ali, Nancy Elgendy, Mohamed Elbadry, Hala Zaid). Author’s contributions: Noha Asem Performed the study design, analysis plan, data management, design data collection tools and performed critical review of the manuscript. Hossam Hosny Massoud planned the study and critical review of the manuscript. Ihab Serag conceptualized the Idea and preparation of convalescent plasma. All other authors contributed equally in study.

## Abstract

**Importance:** Early and effective treatment of COVID-19 is vital for control of SARS-CoV-2 infection

**Objectives:** The primary objective of the study was to assess the degree of clinical improvement in severe and critically ill COVID-19 patients, treated early with early CPT.

**Designs:** An interventional, single-arm, non-randomized clinical trial conducted in Egypt from April 15 to July 21, 2020.

**Settings:** This was a multi-centre study conducted in 3 hospitals in Egypt.

**Participants:** a total of 94 COVID-19 laboratory-confirmed patients using qRT-PCR were enrolled in the study.

**Intervention:** All patients were administered with two plasma units (each unit is 200cc). The volume of donated plasma was 800cc.

**Main Outcome and Measures:** Primary measure was the degree of clinical improvement among the COVID-19 patients who received CPT within seven days

**Results:** A total of 94 patients were enrolled who received CPT either within seven days or after seven days of hospitalization. 82 were severely ill, 12 were critically ill. The average age remained 58 years (±SD 15.1 years). Male were 69% and 49% patients got cured while 51% died with CFR 51%. 75% deaths were above 45years of age. The symptoms were dyspnoea (55%), fever (52%), cough (46%), and loss of taste and smell (21%), and cyanosis (15%). The most common co-morbidities among the <40 years remained Diabetes Mellitus (21%) and Asthma (14%). Among 40-60 years Hypertension (56%), Diabetes Mellitus (39%) and among >60 years age group Hypertension (57%) and Chronic Heart Disease (24%) were reported. CPT within seven days remained significant as compared with the CPT after seven days with the number of days to cure (p=0.007) and ICU stay (P=0.008) among severely ill cured cases.

**Conclusion and Relevance:** Among patients with COVID-19 and severe or critical illness, the use of CPT along with routine standard therapy resulted in a statistically significant improvement when administered within seven days of hospital admission. However, plasma transfusion, irrespective of days to transfusion may not help treat critically ill patients. The overall mean time to cure in severely ill patients was 15 days if CPT provided within seven days with 65% cure rate.

**Trial Registration:** Clinical Intervention identifier: MOHP_COVID-19_Ver1.1

## Introduction

The rapid worldwide spread of the Severe Acute Respiratory Syndrome Coronavirus-2 (SARS-CoV-2) or COVID-19 pandemic from its epicentre; Wuhan has led to an epidemiological breakdown. The virus has caused an array of symptoms predominantly upper respiratory tract infections, and a significant increase in hospitalizations for pneumonia, deaths with multi-organ failure while triggering an economic downturn upending the global economy. A total of51,28 million cases have been reported across 213 countries with 36,34million recovered, and 1.28 million deaths^1^.

Egypt reported its first COVID-19 case on Feb 14, 2020^2,3^. Thereafter, Egypt scaled-up preventive measures, with a partial lockdown starting on March 25. Quantitative real-time polymerase chain reaction (qRT-PCR) was performed extensively for symptomatic patients while patients with a high rate of suspicion, the test was repeated after 48 hours. The current treatment for COVID-19is generally supportive. Several therapeutic agents along with Convalescent Plasma Transfusion are under investigation and data from Convalescent Plasma Transfusions (CPT) have been receiving a lot of attention, after Emergency approvals from the Food and Drug Administration (FDA) suggesting that it may provide a clinical effect in the treatment of SARS-COV-2^4^.

Passive immunization therapies treating infectious diseases date back to the 1920s’, providing instantly available therapeutic strategies for conferring direct immunity to susceptible individuals. Blood from an individual who has recovered from the disease is drawn and screened for high titters neutralizing antibodies, convalescent plasma containing these neutralizing antibodies is then administered in individuals with the infectious disease. Antibodies bind to the pathogen and directly neutralize its effectivity via antibody-mediated pathways, such as complement activation, antibody-dependent cellular cytotoxicity, and/or phagocytosis. Non-neutralizing antibodies that bind to the pathogen contribute to reducing symptoms and mortality, as they do not interfere in the pathogen’s ability to replicate within in-vitro symptoms^5^.

The use of CPT against coronaviruses was first tested during SARS1 outbreak in 2003. One of the largest SARS1 studies involved a total of 80 patients in Hong Kong. Results showed that patients treated before 14 days, had improved outcomes^6^. Eventually, convalescent plasma became established as an empirical treatment during the outbreaks of influenza A (H1N1), where treatment of severe infection with convalescent plasma (n = 20 patients) as associated with reduced respiratory tract, viral load, serum cytokine response, and mortality. Treatment protocols with convalescent plasma became further established for Ebola virus in 2014, and Middle East Respiratory Syndrome (MERS) in 2015^7^.

Clinical trial data from previous studies suggest that using similar CPT protocols could be beneficial in patients infected with SARS-CoV-2. However, there is a lack of robust evidence, generated from the data of large clinical trials that confirm the efficacy of this treatment modality.

The objective of the study was to assess the clinical efficacy of early start of convalescent plasma amongSARS-CoV-2patients as well as to observe intensive care unit (ICU) characteristics and fatality among severe and critically ill patients diagnosed with SARS-CoV-2.

## Methodology

An interventional, single-arm, non-randomized, multi-centre clinical trial was conducted from 15^th^ April to 21^st^ July 2020 in Egypt. The study was approved by the Ministry of Health and Population Ethics Committees (MOHP-EC) and was registered in Egypt no. <u>MOHP_COVID-19_Ver1.1</u>.

Initially, 30 patients were enrolled and later extended where a total of 102 patients tested positive, using quantitative reverse transcriptase-polymerase chain reaction (qRT-PCR) in Central laboratories and upon their admission to the hospital either with severe or life-threatening disease they were included in the study. A total of 94 patients were finally enrolled in the study after exclusion of patients who did not meet the criteria and written informed consent was taken. All patients included in the study have laboratory proven as SARS-CoV-2& had bilateral infiltrates>50% and categorized as severely ill when having hypoxia and critically ill in case of mechanical ventilation use. Patients had received a combination of antiviral agents, antibiotics, and steroids according to the SARS-CoV-2 treatment protocol by Ministry of Health. All patients received 2 convalescent plasma units (Each unit is 200cc)(Figure-1).

**Figure 1:**
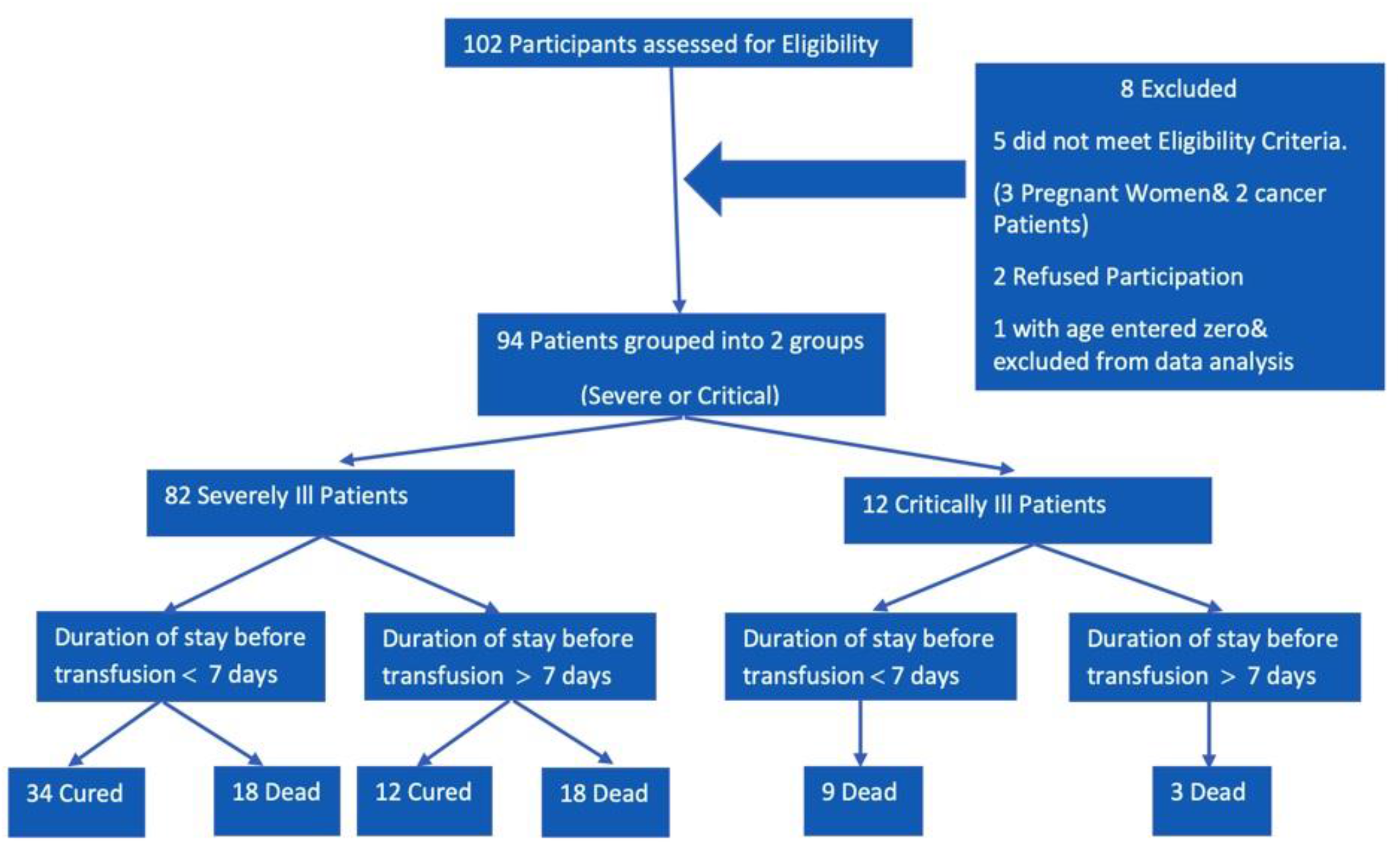
Study Protocol summary

Symptomatic patients were tested for SARS-CoV-2using nasopharyngeal or oropharyngeal swabs through-PCR - a validated assay protocol approved by the World Health Organization (WHO).

### Convalescent plasma donors

Donors between the ages 18-60, with a clinical and laboratory-confirmed recovered diagnosis of SARS-CoV-2were selected. Selected donors had complete resolution of symptoms at least 14 days before donation and negative results for SARS-CoV-2 either from one or more nasopharyngeal swab specimens or by a molecular diagnostic test from the blood. Neutralization antibodies are measured by Microneutralization technique^8^. Donors with a measurement of neutralizing antibody titers more than 1:320 (accepted till 1:80) along with anFDA approved defined serum SARS-CoV-2 specific chemiluminescence antibody titer higher than 1:320 were included. The investigators tested donor units using the Ortho VITROS SARS-CoV-2 total antibodies as per approved FDA guidelines^9^. Units containing anti-SARS-CoV-2 antibodies but not qualified as high titer by the Ortho VITROS SARS-CoV-2 total antibody are considered low titer units.

The volume of donated plasma was 800cc and all donors’ samples were routinely virology screened for HBsAg, HCV Ab, HIV Ag/Ab, Syphilis Ab by using Chemiluminescence technique and were also screened by NAT for HBV DNA, HCV RNA& HIV ½ RNA. All donors provided consent and container labels of convalescent plasma units included the following statement, “Caution: New Drug--Limited to investigational use.”

### Statistical Analysis

Descriptive and inferential analyses were performed. For the quantitative variables; frequencies, averages and standard deviations were calculated. For the timing of the convalescent plasma administered variable coding was done as 1 for CP administration within 7 days of hospitalization and 2 for CP administration after 7 days of hospital administration. Independent t test was applied for comparing the mean duration to cure and ICU stay among the categories of the timing of CP administration. Level of significant was selected as 0.05 and significance values (p) compared. N-1 chi-square test was also applied. IBM® SPSS® was used for analysis.

## Results

### Patient Demographics & Clinical Characteristics

A total of 94 COVID-19 patients including 82 (87%) severely ill and 12 (13%) critically ill were enrolled in the current study and administered two units of convalescent plasma transfusions, within 24 hours, either within 7 days or after 7 days of hospitalization. Patients were divided into three groups; less than 40 years, 40-60 years, and greater than 60 years of age. Each group included 13 (16%), 30 (37%), and 39 (47%) severely ill patients while 1 (8%), 4 (33%), and 7 (58%) critically ill patients, respectively.

The average age remained 58 years (±SD 15.1 years) Among the 94 enrolled patients while the average age among <40 years, 40-60 years and >60 years remained 32(±SD 4), 51 (±SD 6) and 71 (±SD 6). Among the total patients 65 (69%) were male. A total of 46 (49%) patients got cured while 48 (51%) died. The symptoms were dyspnoea (55%), fever (52%), cough (46%), and loss of taste and smell (21%), and cyanosis (15%). The most common co-morbidities among the <40 years remained Diabetes Mellitus (21%) and Asthma (14%). Among 40-60 years age group Hypertension (56%), Diabetes Mellitus (39%), and among >60 years age group Hypertension (57%), Chronic Heart Disease [(24%), and Chronic Kidney Disease were the most frequently reported co-morbidities.

### Severely Ill Cases

Among the total 94 cases 82 (87%) were severely ill cases out of them 46 (56%) cured while 36 (44%) died. A total of 52 (63%) received CPT within seven days of hospital admission while 30 (37%) cases received CPT after seven days. The average days to cure remained 15 days (±SD7.3) among those who received CPT within seven days while the average days to cure remained 23 (±SD 9.4) who received CPT after seven days and difference remained statistically significant p=0.007.

Among the severely ill cases the cure rate among the cases who received CPT within seven days was 65% while among those who received CPT after seven days remained 40%. The difference remained 25% which is statistically significant (p=0.016).

### Severely Ill-Cured

Among the total 82 severely ill cases46 were categorized as severely ill – cured. The average age of severely-cured patients remained 57.3 years (±SD 15 Years). Male were 33 (72%) and female were 13 (28%). Clinical sign and symptoms remained as dyspnoea 44 (96%), cough 38 (82%), fever 34 (74%), cyanosis 10 (21%) and loss of taste and smell 7 (15%). Reported co-morbidities remained as hypertension 23(50%), Diabetic 13(28%), heart disease 6 (13%), Asthma 5 (11%), COPD 4 (9%), Kidney disease 3 (7%) and Chronic liver disease2 (4%).

Out of the total 45 (97%) patients treated with antibiotics and steroids, while 36, (78%) patients were treated with combination of Hydroxychloroquine and 4 (9%) patients were treated with Oseltamivir and Lopinavir/Ritonavir. The mean lowest SPO2, S/F ratio, PAO2, and P/F ratio reported across all age groups as76.15, 190.37, 49.28, and 143.5 respectively. (Table-1)

**Table 1:** Plasma, ICU & Respiratory Characteristics and complications among the severely ill cured subjects (n=46)

| Characteristics | Total | Age Groups (Years) |  |  |
| --- | --- | --- | --- | --- |
|  |  | <40 | 40-60 | >60 |
| Plasma Characteristics |  |  |  |  |
| • Total no. of Patients n (%) | 46(100) | 7 (15) | 18 (39) | 21 (46) |
| • Plasma Administered within 7 Days n (%) | 34(78) | 6 (86) | 12 (67) | 16 (76) |
| • Mean Days to Cure (SD) | 15 (7) | 13.16(4.5) | 15.44 (4.3) | 15.8 (9.4) |
| • Plasma Administered after 7 Days n(%) | 12(22) | 1 (14) | 6 (33) | 5 (24) |
| • Mean Days to Cure (SD) | 23(9) | 28 | 22.16 (11.63) | 22.6 (8.2) |
| ICU Characteristics |  |  |  |  |
| • Mean duration of ICU Stay before Transfusion (SD) | 4.4 (2.7) | 3.71(2.69) | 5.11 (2.44) | 4.04 (3.08) |
| • Mean duration of ICU Stay after Transfusion (SD) | 8.9(7.8) | 5.85 (4.84) | 9.72 (7.47) | 9.28 (8.85) |
| • Mean duration of Ventilator Stay after Transfusion (SD) | 0.1(0.4) | 0 | 0 | 0.23(0.70) |
| Respiratory Characteristics |  |  |  |  |
| • Lowest SPO2 | 76.1(9.5) | 80.2 (26.76) | 71.6 (22.3) | 69.7 (20.2) |
| • Calculated S/F Ratio | 190.3 (23.9) | 200.5 (31.1) | 194.1 (15.6) | 184.7 (26.38) |
| • Lowest PAO2 | 51.3 (19.5) | 67.6 (46.7) | 52.6 (8.6) | 45.1 (17.2) |
| • Lowest P/F Ratio | 143.5(61.6) | 147.3 (45.6) | 132.6 (55.5) | 147.8 (72.54) |
| Complications |  |  |  |  |
| • AKD n(%) | 3 (7%) | 0 (0) | 0 (0) | 3 (14) |
| Pharmacological Treatment |  |  |  |  |
| • Antibiotics | 45 (98) | 7 (100) | 18 (100) | 20 (95) |
| • Oseltamivir | 4 (9) | 1 (14) | 1 (6) | 2 (10) |
| • Hydroxychloroquine | 36 (78) | 5 (71) | 13 (72) | 18 (86) |
| • Lopinavir/Ritonavir | 4 (9) | 0 (0) | 1(6 ) | 3 (14) |
| • Steroids | 45 (98) | 7 (100) | 18 (100) | 20 (95) |

Among the 46 severely-ill cured cases 34 (74%) received plasma within 7 days while 12 (40%) cured cases received plasma after seven days. The average days to cure the cases who received plasma within seven days remained 15 days (± SD 7 days) while the average days to cure who received plasma after seven days remained 23 (±SD 9). The mean difference remained significantly different among both groups (P=0.007). Similarly, the cases who received plasma within seven days the average ICU stay remained 6 days (±SD 4 days) while the cases who received plasma after seven days the average ICU stay was 11 days (±SD 7 days). The difference in the average ICU stay remained significant (p=0.008) (Table 2).

**Table 2:**
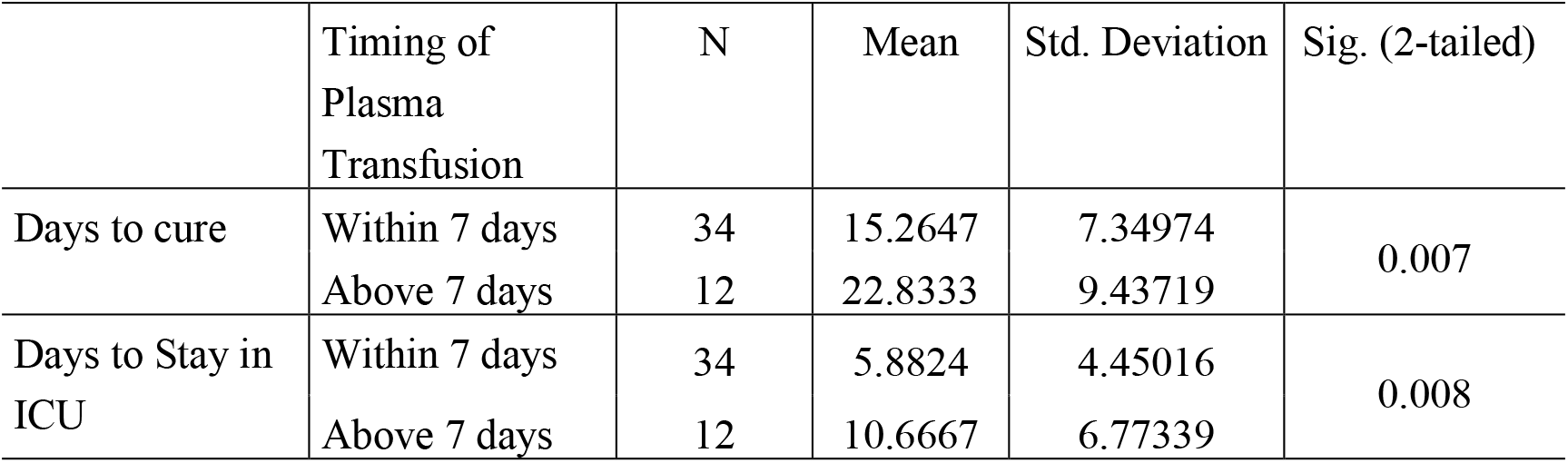
Comparison of Mean days to cure and ICU Stay among cases with timing of Plasma administration (n=46)

### Severely Ill-Dead

Among the total 82 severely ill cases of 36 s were categorized as severely ill - died, patients across all age groups. The average age remained 59 years (±SD 17 Years). Male were 22 (61%) and female were 14 (39 %). Clinical sign and symptoms remained as dyspnoea 33 (92%), cough 30 (83%), fever 29 (81%), cyanosis 14 (39 %) and loss of taste and smell 7 (19%). Reported co-morbidities remained as hypertension 17(47%), Diabetic 11(31 %), heart disease 5 (14%) and Kidney disease 3 (8%). All patients (n=36, 100%) treated with antibiotics and 35 patients (97.2 %) treated with steroids. A combination of Hydroxychloroquine was administered to 69% (n=25) of the subjects and Lopinavir/Ritonavir was given to 11% (n=4) of the subjects. The mean lowest SPO2, S/F ratio, PAO2, and P/F ratio reported across all age groups were 68.96, 155, 56.67, and 129.8, respectively (Table 3). At admission, 32 of the 36 patients reported to the hospital with dyspnoea, which increased progressively before transfusion.

**Table 3:** Plasma, ICU& Respiratory Characteristics and complications among the severely ill Died subjects (n=36)

| Characteristics | Total | Age Groups |  |  |
| --- | --- | --- | --- | --- |
|  |  | <40 | 40-60 | >60 |
| Plasma Characteristics |  |  |  |  |
| • Total no. of Patients n(%) | 36(100) | 6 (17) | 12 (33) | 18 (50) |
| • Plasma Administered within 7 Days n (%) | 18(50) | 2 (33) | 9 (75) | 7 (39) |
| • Mean Days to Die (SD) | 12.5(6.6) | 12 (8.4) | 14.4 (7.7) | 10.1 (4.4) |
| • Plasma Administered after 7 Days n(%) | 18(50) | 4 (67) | 3 (25) | 11 (61) |
| • Mean Days to Die (SD) | 16(7.1) | 16 (7.11) | 13.6 (8.6) | 16.7 (7.4) |
| ICU Characteristics |  |  |  |  |
| • Mean duration of ICU Stay before Transfusion (SD) | 5.4(3.6) | 4.1 (2.6) | 4.9 (3.1) | 6.3 (3.9) |
| • Mean duration of ICU Stay after Transfusion (SD) | 6.6(6.8) | 6.8 (9.7) | 8.5 (7.1) | 5.5 (5.6) |
| • Mean duration of Ventilator Stay after Transfusion (SD) | 6(7.6) | 3.2 (3.8) | 5 (7.3) | 8.5 (9) |
| Respiratory Characteristics |  |  |  |  |
| • Lowest SPO2 (SD) | 65.9(16.2) | 61 (24.3) | 69.2 (16.4) | 66 (14.3) |
| • Calculated S/F Ratio | 165(40.6) | 152.5 (60.9) | 173 (41.1) | 143.6 (59.7) |
| • Lowest PAO2 (SD) | 53.1(16) | 52.00 (22.4) | 53.5 (19.3) | 53.3 (11.8) |
| • Lowest P/F Ratio | 130(51) | 121.5 (111.0) | 150.5 (46.9) | 108.3 (31.8) |
| Complications |  |  |  |  |
| • AKD n(%) | 3(8) | 0 (0) | 1 (8) | 2 (11) |
| **Pharmacological Treatment** |  |  |  |  |
| • Antibiotics | 34 (95) | 6 (100) | 11 (92) | 17 (94) |
| • Hydroxychloroquine | 25 (69) | 4 (67) | 10 (83) | 11 (61) |
| • Lopinavir/Ritonavir | 4 (11) | 0 (0) | 3(25 ) | 1 (8) |
| • Steroids | 35 (97) | 6 (100) | 12 (100) | 17 (94) |

Among the severely ill died cases 18 (50%) received plasma within 7 days while 18 (50%) received plasma after seven days. The cases who received plasma within seven days the average ICU stay remained 7 days (±SD 5 days) while the cases who received plasma after seven days the average ICU stay was 4 days (±SD 3 days). The difference remained significant (p=0.04) (Table 4).

**Table 4:** Comparison of Mean days of ICU Stay among cases with timing of Plasma administration among severely ill-died cases(n=36)

|  | Timing of Plasma Administration | N | Mean | Std. Deviation | Std. Error Mean |
| --- | --- | --- | --- | --- | --- |
| Days of ICU Stay | Within 7 days | 18 | 6.8333 | 5.09036 | 0.04 |
|  | Above 7 days | 18 | 3.8333 | 3.45134 |  |

### Critically Ill-Dead

Out of the total 12 patients were categorized as critically ill - died, patients across all age groups. The average age remained 57 years (±SD 12 Years). Male were 10 (83%) and female were 2 (17 %). Clinical sign and symptoms remained as dyspnoea 12 (100%), cough& Fever9 (75%), cyanosis 6 (50 %) and loss of taste and smell 2 (17 %). Reported co morbidities remained as hypertension 6 (50 %), Diabetic 4 (33 %), heart disease, Kidney diseases and Asthma remained2 (17%).

All the critically ill died cases were treated with steroid while 9 (75%) cases were treated with antibiotics, a combination of Hydroxychloroquine was given to 10 (83%) and Lopinavir/Ritonavir was provided to 1 (8%) (Table 5).

**Table 5:** Plasma, ICU & Respiratory Characteristics and complications among the critically ill Died subjects (n=12)

| Characteristics | Total | Age Groups |  |  |
| --- | --- | --- | --- | --- |
|  |  | <40 | 40-60 | >60 |
| Plasma Characteristics |  |  |  |  |
| • Total no. of Patients n(%) | 12(100) | 1(8) | 4 (33) | 7 (58) |
| • Plasma Administered within 7 Days n (%) | 9(75) | 1 (100) | 3 (75) | 5 (71) |
| • Mean Days to Die (SD) | 15.2(9.9) | 24 | 7.33 (3) | 20.5 (10.9) |
| • Plasma Administered after 7 Days n(%) | 3(25) | 0 (0) | 1 (25) | 2 (28) |
| • Mean Days to Die (SD) | 26.3(15.1) | 0 | 10 | 34.5 (7.77) |
| ICU Characteristics |  |  |  |  |
| • Mean duration of ICU Stay before Transfusion (SD) | 4.9(2.9) | 6 | 3 (1.1) | 5.8 (2.5) |
| • Mean duration of ICU Stay after Transfusion (SD) | 11.1(11.5) | 18 | 3 (2.6) | 15.2 (12.60) |
| • Mean duration of Ventilator Stay after Transfusion (SD) | 8.3(10.2) | 18 | 3.3 (2.2) | 11.5 (12.8) |
| **Respiratory Characteristics** |  |  |  |  |
| • Lowest SPO2 | 71.7(15) | 60 | 70 | 74 (16.8)) |
| • Calculated S/F Ratio | Not Reported | Not reported | Not reported | Not reported |
| • Lowest PAO2 | 51(9.3) | 50 | 45 | 52.3 (10.6) |
| • Lowest P/F Ratio | 90.3(14.6) | 70 | Not reported | 94.4 (11.9) |
| **Complications** |  |  |  |  |
| • AKD n(%) | 4 (33) | 0(0) | 1(25) | 3 (43) |
| **Pharmacological Treatment** |  |  |  |  |
| • Antibiotics | 9 (75) | 1 (100) | 2 (50) | 6 (86) |
| • Oseltamivir | 0(0) | - | 0 (0) | 0 (0) |
| • Hydroxychloroquine | 10 (83) | 1(100) | 3 (75) | 6 (86) |
| • Lopinavir/Ritonavir | 1 (8) | - | 1(25 ) | 0 (0) |
| • Steroids | 12(100) | 1 (100) | 4 (100) | 7 (100) |

The mean lowest SPO2, PAO2, and P/F ratio reported across all age groups were 71.75, 51.13, and 90.33, respectively. It was observed that there was no significant difference between patients administered with plasma within or after 7 days of administration.

### Case Fatality

Out of the total 94 COVID-19 Cases 46 cases got cured while 48 cases died (CFR 51%). Among the total died cases the average age remained 58 (±SD 15). The most frequent age among died cases was 73 (Mode), 25% deaths were under 45 years of age(First quartile), 50% deaths were up to 61 years of age (Second Quartile), 75% deaths were up to 70 years of age (Third Quartile) and 25% deaths were above the 70 years of age. 67% male were died and 33% female died. Respiratory complications were more reported among the dead cases.

## Discussion

The current study has shown that early administration of convalescent plasma in COVID-19 patients remained a critical measure for its clinical efficacy in severely ill patients. However, plasma transfusion, irrespective of days to transfusion may not be helpful in treating critically ill patients.

In history, several studies^10,11, 12^have reported treatment with convalescent plasma transfusions for viral infections. Similar results were provided by a study published in china on April 2020, a total of 10 severely ill COVID-19 patients were enrolled in the study and single dose of 200ml plasma was administered to each of the subject and there was significant improvement in the clinical symptoms^13^

Some other published studies also provided the plasma transfusion should be done during the early phase of the diseases and will be more beneficial^14, 15^. China provided the usefulness of plasma transfusion among the critically ill COVID-19 cases. In this study 5 critically ill cases were enrolled and received transfusion body temperature normalized within 3 days in 4 of 5 patients, the SOFA score decreased, and PAO2/FIO2 increased within 12 days (range, 172-276 before and 284-366 after). Viral loads also decreased and became negative within 12 days after the transfusion.^16^

Similar results were observed in influenza A (H1N1) patients, where patients who received plasma transfusions (n=20) had significantly fewer deaths when compared to controls (n=73) deaths (20% vs. 54.8%; P=.01).^17^ These positive effects of plasma transfusions in viral infections can be attributed to the humoral immunity established in recovered patients containing a large quantity of neutralizing antibodies capable of neutralizing SARS-CoV-2 and eradicating the pathogen from blood circulation and pulmonary tissues. In this multi-centre non-randomized study, 94 patients (82 severely ill and 12 critically ill) with SARS-CoV-2 were treated with convalescent plasma. As assessed by days to transfusion (before and after 7 days) from date of admission, clinical symptoms such as dyspnoea, cyanosis, fever, and cough improved within days of transfusion of plasma. This response time may have impact on pandemic economy as shorter duration of hospital stay leads to reduced cost. A study was published to examine the economic consequences of hospital admissions. And it is evident that with long stay there is a significant implication of the economics and increase of the out of pocket^18^. The current study provided that clinical respiratory parameters also improved in all cured patients. The results, therefore, support the initial hypothesis stating that CPT at an early stage may be effective in treating SARS-CoV-2. Similar results were provided by a study published during June 2020 in China. This study comprehensively evaluated the effectiveness, safety, and indications of convalescent plasma transfusion (CPT) therapy for severe or critical COVID-19 patients. It is provided that 70% of the cases with severe respiratory symptoms showed improvement and removed oxygen supports within 7 days after CPT19.

Our study also reports the deaths of critically ill patients who were placed on invasive mechanical ventilation before plasma transfusions. All 12 patients in this group died& they had respiratory system complications predominantly including pneumonia. All patients, irrespective of days to plasma transfusions had prolonged ICU stay similar to other studies. Similar results are evident form a meta- analysis published during May 2020 and provided that among the fatal cases the prevalence of respiratory co morbidities is higher [10.89% (95%CI: 7.57%, 15.43%)] as compared to the total cases [3.65% (95%CI: 2.16%, 6.1%)]^20^

In previous studies^21^ viral load was linked to severity of SARS-COV-2. Thereby, supporting the current clinical evidence that suggests, clinical benefit is most likely in patients treated early in the course of the disease during viremia stage. Another study in 2018 was based on viral load and sequence analysis provided that viral load is significantly associated with the severity of diseases^22^.

These results are comparable with other SARS-CoV-2convalescent plasma transfusion studies that have been published since the beginning of this pandemic^23, 24, 25^. Shen et al. conducted one of the first plasma transfusions studies with a small sample size of five patients. The results reported, the PAO2 levels ranged from 172 to 276 before transfusion which was improved among four of five patients within seven days after transfusion (overall range: 206 - 290), and improved substantially (range: 284-366) on the 12th day after the plasma treatment. Chen et al. also reported improvement of clinical symptoms (fever, cough, shortness of breath, and chest pain) in all 10 patients within 1 to 3 days of plasma transfusions.^16^.

### Limitations

Due to the clinical features of included patients; the investigators could not conduct a randomized controlled trial. However, we understand that a controlled study may further help in establishing the dynamics of the viremia of SARS-CoV-2 while capturing the optimal transfusion time point. Additionally, all patients were also treated with multiple pharmaceutical agents including antibiotics, antivirals, and steroids requiring robust comparative analyses to clearly establish the effect of each treatment modality individually and collectively as well.

## Conclusion

This non-randomized clinical trial demonstrates the clinical efficacy of early convalescent plasma in treating SARS-CoV-2 infected patients. The study results indicate that cure rates are significantly better when plasma is administered within the first seven days of hospital admission for severe cases. The overall cumulative mean time to cure remained 15.2 days if CPT provided within seven days with 65% cure rate as compared to 22.8 days if CPT administered after seven days with 40% cure rate. For critically ill patients, the study reports that plasma transfusions may not be beneficial, due to the rapid progression of the infection and associated complications. We recommend well-designed randomized studies to further establish its efficacy in the future.

## Data Availability

all data are available

## Acknowledgment

All clinical coordinators participated in coordinating field work and data entry.

## Conflict of Interest

No conflict of interest

## Notes

### Competing Interest Statement

The authors have declared no competing interest.

### Clinical Trial

ClinicalTrials.gov Identifier: NCT04816942

### Author Declarations

The study was approved by the Ministry of Health and Population Ethics Committees (MOHP-EC) and was registered in Egypt no. MOHP_COVID-19_Ver1.1.

## References

1. COVID-19 Dashboard by the Center for Systems Science and Engineering (CSSE) at Johns Hopkins University, 24th October 2020. https://coronavirus.jhu.edu/map.html

2. Mohammed A Medha. Mohamed El Kassas. COVID-19 in Egypt: Uncovered figures or a different situation?. J Glob Health. 2020 Jun; 10(1): 010368.. doi: 10.7189/jogh.10.010368 Published online 2020 Jun 17. https://www.ncbi.nlm.nih.gov/pmc/articles/PMC7303805/

3. Estimation of COVID-19 burden in Egypt. Thelancet. 2020 Aug. https://doi.org/10.1016/S1473-3099(20)30319-4

4. Chai KL, Valk SJ, Piechotta V, Kimber C, Monsef I, Doree C, Wood EM, Lamikanra AA, Roberts DJ, McQuilten Z, So-Osman C, Estcourt LJ, Skoetz N. Convalescent plasma or hyperimmune immunoglobulin for people with COVID-19: a living systematic review. Cochrane Database of Systematic Reviews 2020, Issue 10. Art. No.: CD013600. DOI: 10.1002/14651858.CD013600.pub3

5. Barney S. Graham, Donna M. Ambrosino. History of Passive Antibody Administration for Prevention and Treatment of Infectious Diseases. CurrOpin HIV AIDS. Author manuscript; available in PMC 2016 May 1.Published in final edited form as: CurrOpin HIV AIDS. 2015 May; 10(3): 129–134. doi: 10.1097/COH.0000000000000154. PMCID: PMC4437582

6. Y. Cheng, R. Wong, Y. O. Y. Soo, W. S. Wong, C. K. Lee, M. H. L. Ng, P. Chan, K. C. Wong, C. B. Leung, G. Cheng. Use of convalescent plasma therapy in SARS patients in Hong Kong.Eur J Clin Microbiol Infect Dis. 2005; 24(1): 44–46. Published online 2004 Dec 23. doi: 10.1007/s10096-004-1271-9.PMCID:PMC7088355

7. Giuseppe Marano, Stefania Vaglio, Simonetta Pupella, Giuseppina Facco, Liviana Catalano, Giancarlo M. Liumbruno, Giuliano Grazzini. Convalescent plasma: new evidence for an old therapeutic tool?.BloodTransfus. 2016 Mar; 14(2): 152–157. Prepublished online 2015 Nov 6. doi: 10.2450/2015.0131-15.PMCID:PMC4781783

8. Fatima Amanat, Kris M. White, Lisa Miorin, Shirin Strohmeier, Meagan McMahon, Philip Meade, Wen-Chun Liu, Randy A. Albrecht, Viviana Simon, Luis Martinez-Sobrido, Thomas Moran, Adolfo García-Sastre, Florian Krammer2 An In Vitro Microneutralization Assay for SARS-CoV-2 Serology and Drug Screening. CurrProtocMicrobiol. 2020 Sep; 58(1): e108. Published online 2020 Jun 25. doi: 10.1002/cpmc.108

9. Ortho Clinical Diagnostics – FDA. https://www.fda.gov/media/136967/download

10. Research C for BE, Investigational. COVID-19 convalescent plasmademergency INDs. Available at: FDA; 2020. http://www.fda.gov/vaccinesblood-biologics/investigational-new-drug-ind-or-device-exemption-ideprocess-cber/investigational-covid-19-convalescent-plasma-emergency-inds. [Accessed 15 May 2020].

11. NIH. US National Library of Medicine. Available at: https://clinicaltrials.gov/ct2/results?cond¼COVID-19&term¼randomizedþconvalescentþplasma&cntry¼&state¼&city¼&dist¼&Search¼Search&type¼Intr. [Accessed 15 May 2020].

12. Bloch EM, Shoham S, Casadevall A, Sachais BS, Shaz B, Winters JL, et al. Deployment of convalescent plasma for the prevention and treatment of COVID-19. Available at: J Clin Invest 2020. http://www.jci.org/articles/view/138745. [Accessed 15 May 2020]

13. Kai Duan, Bende Liu, Cesheng Li, View ORCID Profile Huajun Zhang, Ting Yu, View ORCID ProfileJieming Qu, View ORCID ProfileMin Zhou, View ORCID ProfileLi Chen, Shengli Meng, Yong Hu, Cheng Peng, Mingchao Yuan, View ORCID Profile Jinyan Huang, Zejun Wang, Jianhong Yu, Xiaoxiao Gao, Dan Wang, View ORCID ProfileXiaoqi Yu, View ORCID ProfileLi Li, Jiayou Zhang, Xiao Wu, Bei Li, View ORCID ProfileYanping Xu, Wei Chen, Yan Peng, Yeqin Hu, Lianzhen Lin, Xuefei Liu, Shihe Huang, Zhijun Zhou, Lianghao Zhang, Yue Wang, Zhi Zhang, Kun Deng, Zhiwu Xia, Qin Gong, Wei Zhang, Xiaobei Zheng, Ying Liu, Huichuan Yang, Dongbo Zhou, Ding Yu, Jifeng Hou, Zhengli Shi, Saijuan Chen, Zhu Chen, Xinxin Zhang, and View ORCID ProfileXiaoming Yang. Effectiveness of convalescent plasma therapy in severe COVID-19 patients. PNAS April 28, 2020 117 (17) 9490–9496; first published April 6, 2020; https://doi.org/10.1073/pnas.2004168117

14. World Health Organization. Use of Convalescent whole blood or plasma collected from patients recovered from Ebola virus disease for transfusion, as an empirical treatment during outbreaks. Interim Guidance Natl Health Authorities Blood TransfusServ 2014;1:1e19.

15. Hegerova L, Gooley T, Sweerus KA, Maree CL, Bailey N, Bailey M, et al. Use of convalescent plasma in hospitalized patients with Covid-19dcase series. Blood 2020:2020006964.

16. Shen C, Wang Z, Zhao F. Treatment of 5 Critically Ill Patients With COVID-19 With Convalescent Plasma. JAMA. 2020;323(16):1582–1589. doi:10.1001/jama.2020.4783

17. Hung IF, To KK, Lee CK, et al. Convalescent plasma treatment reduced mortality in patients with severe pandemic influenza A (H1N1) 2009 virus infection. Clin Infect Dis. 2011;52(4):447–456

18. Carlos Dobkin, Amy Finkelstein, Raymond Kluender, Matthew J. Notowidigdo.The Economic Consequences of Hospital Admissions.Am Econ Rev. Author manuscript; available in PMC 2018 Feb 12.Published in final edited form as: Am Econ Rev. 2018 Feb; 102(2): 308–352. doi: 10.1257/aer.20161038.PMCID: PMC5809140

19. Xinyi Xia, Li Kening, Lingxiang Wu. Improved Clinical Symptoms and Mortality on Severe/Critical COVID-19 Patients Utilizing Convalescent Plasma Transfusion. Blood 136(6) DOI: 10.1182/blood.2020007079

20. Morgan Spencer Gold, Daniel Sehayek, SofianneGabrielli, Xun Zhang, Christine McCusker& Moshe Ben-Shoshan (2020): COVID-19 and comorbidities: a systematic review and meta-analysis, Postgraduate Medicine, DOI: 10.1080/00325481.2020.1786964

21. Heneghan, Carl, Brassey, Jon, Jefferson, Tom. SARS-CoV-2 viral load and the severity of COVID-19. March 2020.https://www.cebm.net/covid-19/sars-cov-2-viral-load-and-the-severity-of-covid-19/

22. Ng KT, Oong XY, Lim SH, et al. Viral load and sequence analysis reveal the symptom severity, diversity, and transmission clusters of rhinovirus infections. Clin Infect Dis. 2018;67(2):261–268.

23. van Pottelberge G.R, Vlasveld I.N, Ammerlaan H.S.M. Convalescent plasma for COVID-19: a randomized clinical trial.medRxiv. 2020; https://doi.org/10.1101/2020.07.01.20139857

24. Eckhardt C.M., Cummings M.J, Rajagopalan K.N., Borden S. Evaluating the efficacy and safety of human anti-SARS-CoV-2 convalescent plasma in severely ill adults with COVID-19: a structured summary of a study protocol for a randomized controlled trial.Trials. 2020; 21:499

25. Hegerova L., Gooley T., Sweerus K.A., Maree C.L.Use of convalescent plasma in hospitalized patients with Covid-19: case series. Blood. 2020; 136:759–762

